# Ethics in medical research: A quantitative analysis of the observations of Ethics Committees in research protocols

**DOI:** 10.1101/2024.06.23.24309373

**Authors:** Santiago Vasco-Morales, Gabriel Alejandro Vasco-Toapanta, Cristhian Santiago Vasco-Toapanta, Paola Toapanta-Pinta

## Abstract

**Objective:** To determine the frequency of observations made by Research Ethics Committees (RECs) regarding non-compliance with ethical principles in research.

**Methods:** We searched for articles published up to November 30, 2023. In the databases: PubMed, Scopus and Google Scholar. Single-proportion meta-analyses were performed with the R V.3.6.1 program.

**PROSPERO Registry:** CRD42021291893

**Results:** 9 publications were reviewed, including cross-sectional, retrospective cohort, and descriptive studies. Lack of adherence to the ethical principle of justice was detected in up to 100% of the protocols evaluated. In addition, 9% (95% CI: 7-12) of observations in Latin America and 15% (95% CI: 9-24) in Europe. Autonomy was observed in 26% (95% CI: 20-33) of the protocols, reaching 17% (95% CI: 13-22) in experimental studies. Beneficence, lack of adherence in the protocols evaluated from 41.17% to 77.38%, observations per protocol ranged from 5.26% to 27.11%.

**Discussion:** The findings highlighted disparities between regions and types of studies, reflecting cultural, interpretive, and human and institutional resource differences. RECs should ensure thorough and equitable assessments, promote fair selection, respect autonomy, and maximize benefits while minimizing risks to participants.

This study provides an assessment of ethical practices in medical research, highlighting key areas for improving compliance with fundamental ethical principles.

## Introduction

Throughout history, multiple events have evidenced the mistreatment and abuse of human beings in the name of knowledge and scientific advances. These abuses have forced society to establish guidelines and norms that prioritize respect for life and the integrity of people. In this context, ethical requirements have been structured that are currently indispensable for scientific research.(1)

Among the most representative documents that refer to ethical principles are:

1) The Nuremberg Code created in 1947 by an international court, where the first and longest of its principles establishes the voluntary informed consent of the individual as essential and indicates that the objective of research must be directed to the common good of society.(2)
2) The Declaration of Helsinki in 1964, developed by the World Medical Association, which included guidelines for research on humans who lack decision-making power and vulnerable groups.
3) The Belmont Report carried out in 1979 by the National Commission for the Protection of Human Subjects identified three basic ethical aspects for all research: a) Respect for people or autonomy through which the patient has the freedom to choose and the power to decide whether or not to participate in the research, in addition, protection for those who are incapable of deciding for themselves and the obtaining of informed consent after understanding, understanding and voluntary participation was included, b) Beneficence, which means maximizing benefits or seeking the well-being of the patient, protecting his life, health, privacy and dignity. It is accompanied by the non-maleficence or the moral obligation of the researcher not to harm or minimize it, for which it is essential that in addition to analyzing the risk/benefit ratio of the procedures to be used, he is responsible for the consequences and not abandons the patient and c) the justice in which the ethical requirements are met, legal and legal requirements of each country so that the results or benefits of the research are shared equitably among the population groups, therefore this principle directly influences the selection of the research subjects(3).

It is indisputable that medical practice needs scientific research for the generation of new knowledge, in which experimentation on human subjects is required, the same that is currently governed by the ethical principles published in the different international consensus, therefore within the laws of medical research, to guarantee compliance with these principles, the Research Ethics Committee (CEI) has been created in different institutions Within the countries, which in addition to safeguarding the rights, dignity, safety and well-being of the participants, must offer the public guarantee and compliance with the methodological steps of the research, for which a series of forms have been developed and established that the researcher must complete and include in the presentation of the protocol. This situation has led to a progressive increase in the proportion of research protocols submitted and requiring modifications(4,5).

Within the methodology of a protocol to comply with ethical principles, when the research question is posed, it must also be questioned whether the results to be found will benefit the participants and whether they will provide new knowledge, in the event that no benefits are expected for the participants or they are exposed to risks, and there are no contributions to knowledge, the principles of beneficence are violated; but if there are also resources available to carry out studies of little or no relevance, the principle of justice is not complied with, by not allowing these resources to be used for other important needs of the population, therefore justice is constituted as a social principle. In other words, in order to comply with the principles of beneficence and non-maleficence, the researcher must be diligent and careful in the choice of the type of study, the sample, the objectives and their deadline for compliance, as well as the protection of the integrity of the participants, the proportion between risks and benefits, the preservation of privacy, etc. the manipulation of results and their objective analysis(6,7).

In recent years, the Informed Consent Form has become a prerequisite for people to participate in research and consists of a document through which the researcher requests a mentally competent patient’s approval to perform a procedure, after the due explanation of both the methodology, the methodology and the the objectives, effects, risks and benefits of the same; For this explanation to be adequate, simple, clear and understandable language must be used, avoiding bias and without exercising coercion, only in this way is autonomy respected and the protection of rights such as confidentiality and the duties of the participants is ensured. Only the Ethics Committee can authorize the completion of the informed consent form in case of more than minimal risk(8)

The ethics committees consider the following to be breaches of the principle of autonomy: the absence of the informed consent request document, the lack of specificity in its wording, the lack of information on the subject’s participation, the absence of consent for the storage and destination of the data, clarification on the type of evidence and its repercussions, as well as the absence of a guarantee of the principle of confidentiality, the absence of the right to know the results of the research and the lack of adequate description of the procedures that the participant must follow to exercise their rights such as access, rectification, cancellation or opposition. Among the failures to comply with the principles of beneficence and non-maleficence, the following are named: the conduct of research on subject’s incapable of providing informed consent for themselves and vulnerable people, the lack of priority of the diagnostic-care interest over the interests of the research, the failure to handle coded or anonymized samples, and the failure to ensure harm (4,5,9).

The objective of this study was to determine the frequency of observations made by Research Ethics Committees regarding non-compliance with ethical principles in medical research. Additionally, we sought to analyze the temporal evolution of these observations, identify possible geographical variations in their frequency, and determine if the type of study influenced the number of observations received.

## Material and Method

The protocol for this study is in the International Prospective Registry of Systematic Reviews PROSPERO under code CRD42021291893.

### Search Strategy

The search was carried out in databases available online and published until November 30, 2023. To search for the articles, the following databases were examined: PubMed, Scopus and Google Scholar, using the keywords: “Ethics Committees, Research”, “Ethics Committees”, “Committee, Research Ethics”, “Committees, Research Ethics”, “Boards, Institutional Review”, “Institutional Review Boards”, “Review Boards, Institutional”, “Review Board, Institutional”, “IRB”, “Research Ethics Committees”, “IRBs”, “Research Ethics Committee”, “Ethics Committee, Research”, “Board, Institutional Review”, “Institutional Review Board”, “Principle-Based Ethics”, “Principle-Based Ethics”, “Clinical Protocols”, OR “Clinical Trial Protocol”, “Beneficence”, “Personal Autonomy”, “Justice”, “Benevolence”, “Nonmaleficence”, “Personal Autonomy”, “Autonomy, Personal”, “Self Determination”, without language restriction. The search equations were performed by combining the terms MeSH with the Boolean operators AND and OR.

### Procedure for searching, extracting and analysing data

C.V carried out the bibliography search in the databases. S.V, PT and CV independently selected the articles according to the title and reading of the full text, and the inclusion and exclusion criteria were applied to decide which articles were included in the study. When there was disagreement, all the authors in consensus decided. The extraction of the data from the tables or the body of the text and conversion to the base format in a spreadsheet was performed by GV. SV performed the meta-analyses. The final collation of the data in the tables and results of the statistical analysis was carried out by PT, the interpretation of the results and the discussion was carried out by SV and CV, all authors contributed to the writing of the text and approved the final version.

### Statistical method

To perform a single-ratio meta-analysis, the number of observations and sample size of each included study were extracted. The statistical program R version 4.2.2 (2022) was used together with the “meta” package, to calculate an overall proportion using the inverse variance method and the generalized linear mixed model (GLMM). To assess heterogeneity, we applied the interpretation criteria described in the Cochrane Handbook. The input data corresponded to sample sizes (number of observations related to non-compliance with ethical principles) and the total number of protocols analysed or observations made. The number of observations in the reports that provided only the number of protocols evaluated along with the proportion of the observations was made by the Ombudsman(10,11)

To address and present the results appropriately, the following were considered:

- The specific characteristics of the studies
- The application of the random-effects model, which assumes that the included studies represent a random sample of the universe of possibilities, especially when the number of participants varies considerably between studies.

Given the nature of the study, significant heterogeneity was anticipated, so subgroup analyses were also considered according to:

- The year of publication of the study
- Whether the Research Ethics Committees (RECs) exclusively analyzed experimental studies (clinical trials)
- The geographical region (America or Europe)

To assess publication bias, we used Egger’s linear regression, adjusted for application with a minimum of three studies. A p< value of 0.1 was considered to suggest the presence of bias.

### Inclusion criteria, study quality, risk of bias and quality of evidence. Inclusion criteria

Observational studies, including retrospective, cross-sectional, cohort, and analytical descriptive designs.

Studies that investigated the activity and functioning of Research Ethics Committees (RECs).

Studies that provided quantitative and/or qualitative data on the ethical review process of research protocols by RECs.

Studies that reported indicators related to the evaluation of ethical principles in research, such as: number of observations made, number and type of ethical recommendations.

### Exclusion Criteria

Studies in which the objective did not correspond to the research question. We excluded studies focusing on:

- The operation of healthcare ethics committees or other committees not related to the ethical review of research.
- Ethical aspects related exclusively to animal studies.

The constitution, distribution or characteristics of the staff of the ethics committees, without addressing their ethical review activity.

- Narrative reviews or other types of documents that did not present original data.

Opinion articles, editorials, letters or any other format without providing data.

Exclusion Criteria

Studies that are not available in full text.

### Quality of studies

The critical evaluation of the included studies was carried out with the tools of the Jhoana Brighs Institute: 1) for case series, 2) for cross-sectional studies and 3) for cohort studies(12,13).

### Quality of the evidence found

To assess the quality of the evidence found, we applied the criteria of the GRADE manual, which classifies the certainty of the evidence into four levels: very low, low, moderate and high.

## Results

### 1. Inclusion of studies

Nine publications were identified that described the variables of interest in the research protocols submitted to the RECs. The study selection process is represented in the flowchart presented in Figure 1.

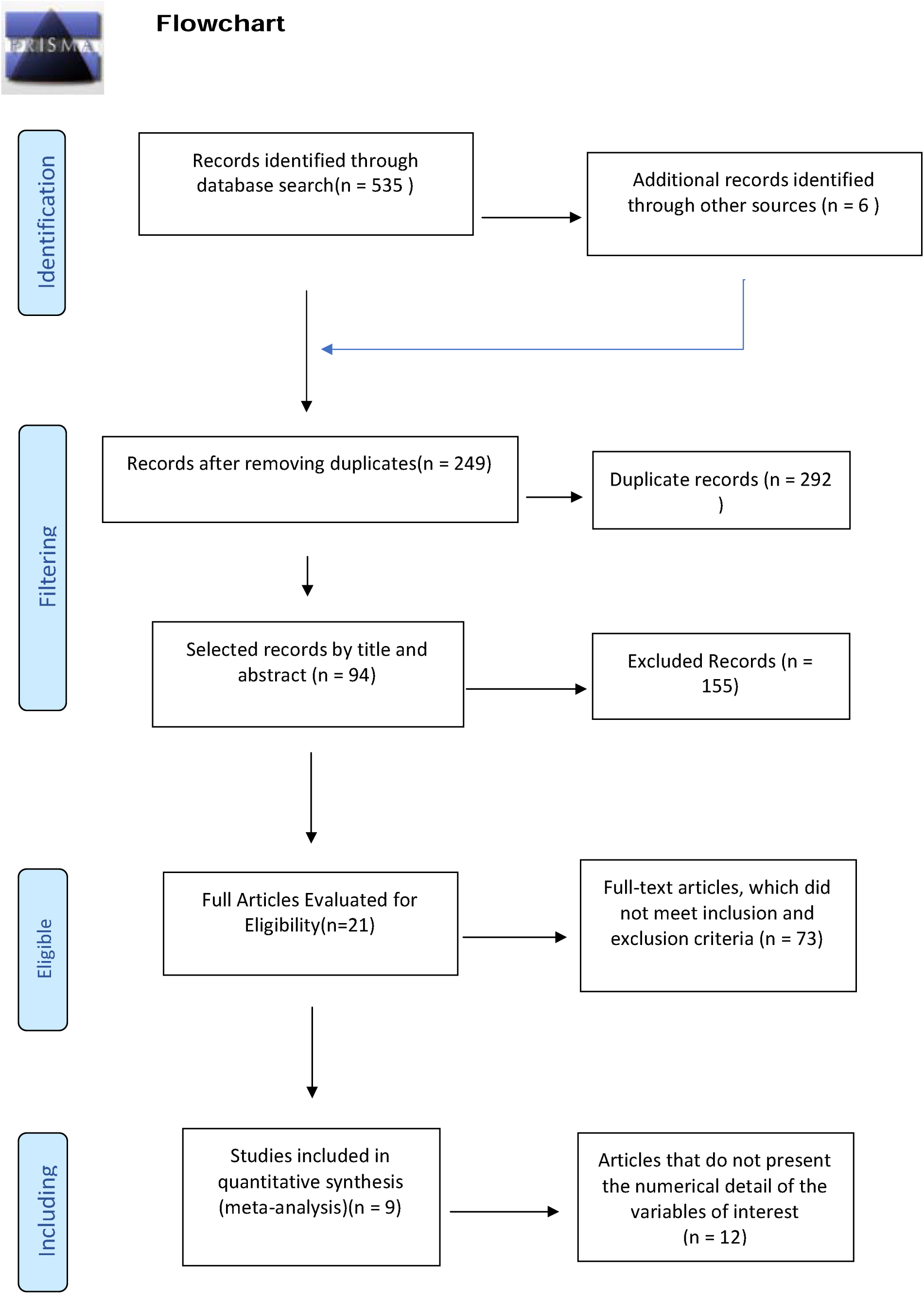

The overview of the included studies is presented in Table No.1

**Table 1.** Description of selected studies.

| **Author** | **Year** | **Protocols**<br>**evaluated** | **Type of stu-**<br>**dy** | **Country** | **Critical**<br>**appraisal** |
| --- | --- | --- | --- | --- | --- |
| Allen P | 1982 | 751 | Descriptive | England | 77,77 |
| Dal-Re R | 2004 | 183 | Descriptive | Spain | 83,33 |
| Decullier E | 2005 | 976 | Retrospective<br>cohort | France | 88,88 |
| Minaya-Martinez G | 2008 | 91 | Transverse | Peru | 77,7 |
| Well, M | 2009 | 357 | Transverse | Brazil | 88,88 |
| Martín-Arribas, M. | 2012 | 66 | Descriptive | Spain | 83,33 |
| Gonorazky, S. E * | 2008 | 33 | Descriptive | Argentina | 83,33 |
| Castañeda Iñiguez<br>M * | 2014 | 51 | Descriptive | Chile | 72,22 |
| Torres Cornejo K * | 2017 | 84 | Retrospective<br>cohort | Peru | 83,33 |

\* These studies describe the observations made according to the number of protocols evaluated.

The summary of the data on the observations of informed consent and ethical principles is shown in Table No.2.

**Table 2.**
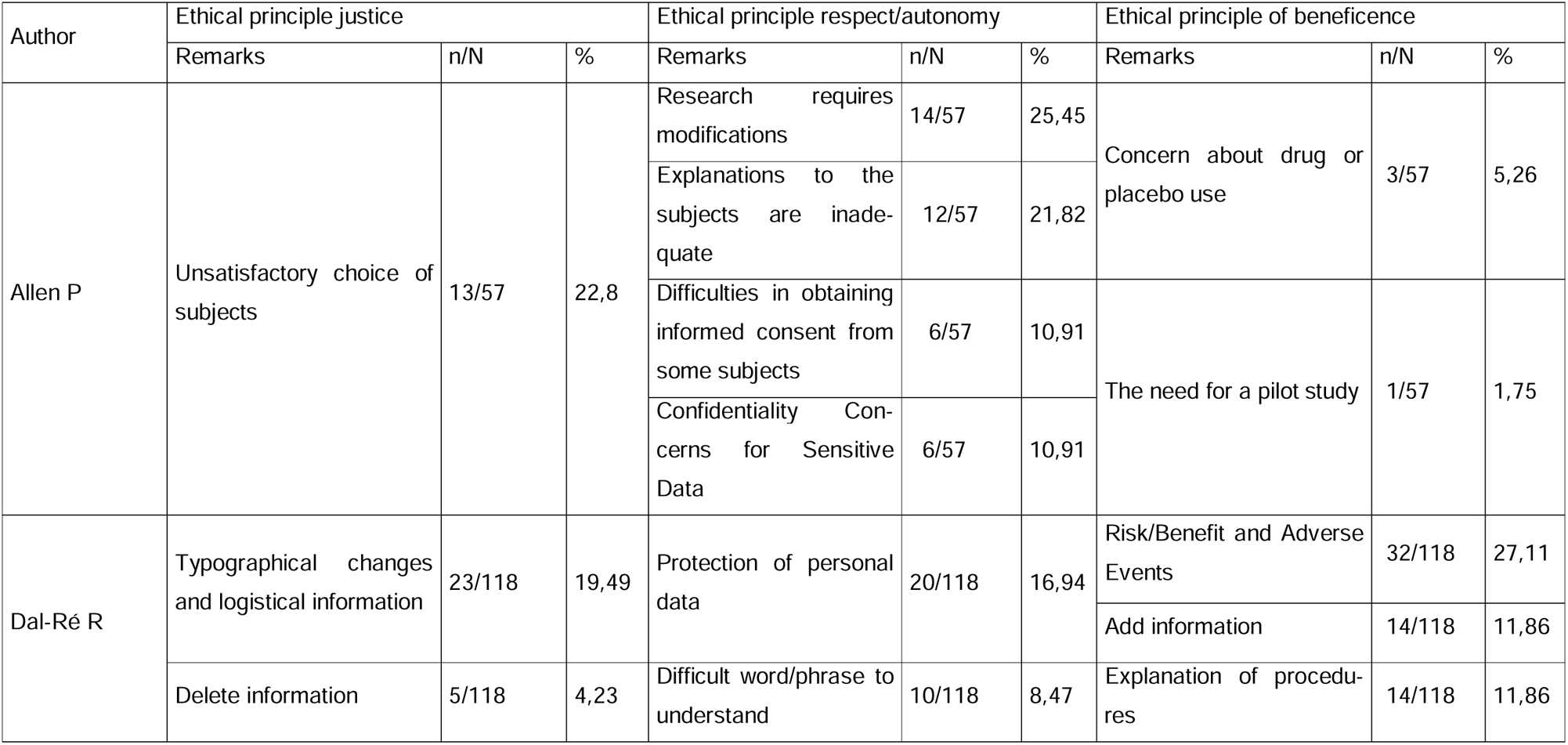
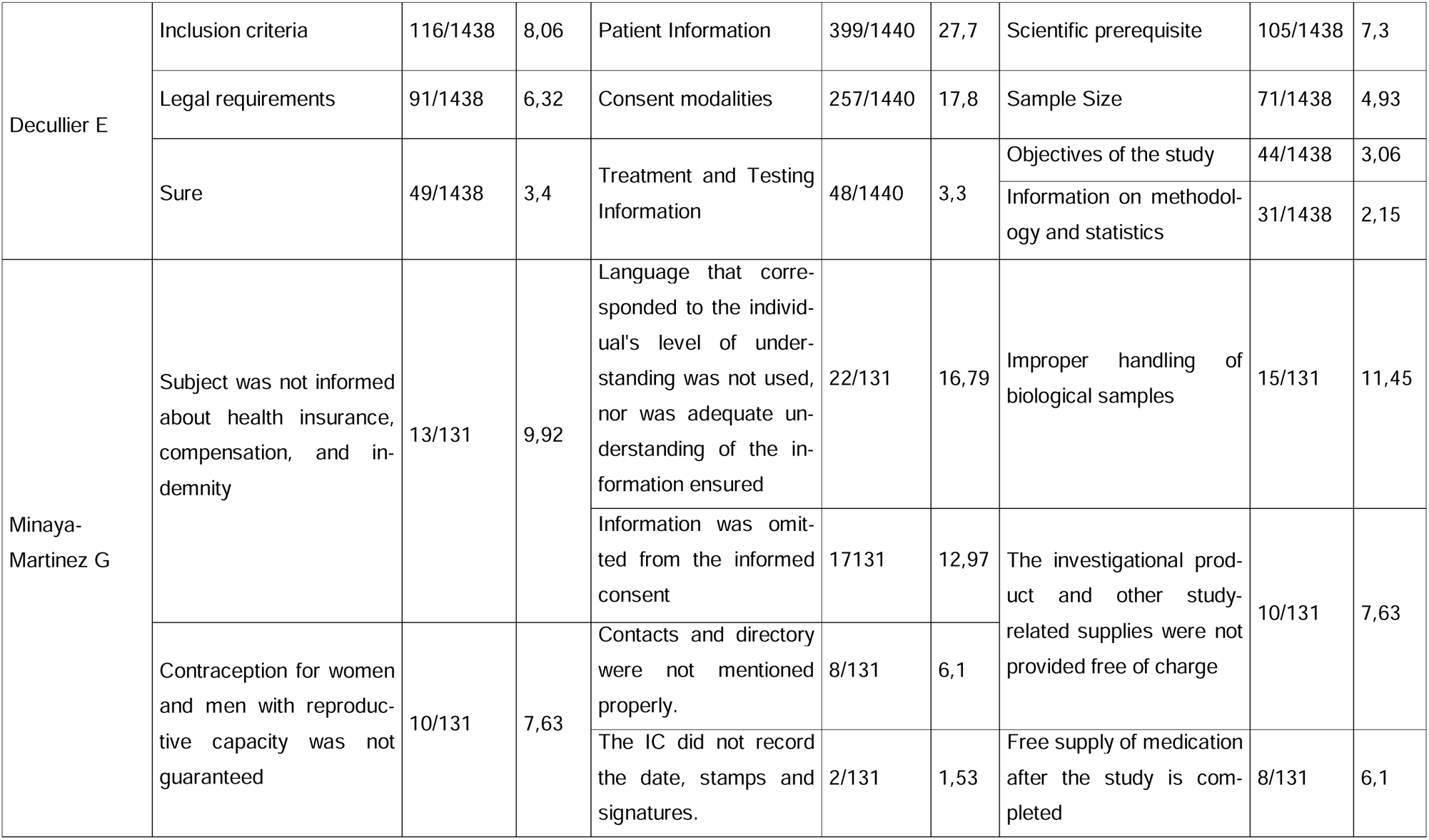
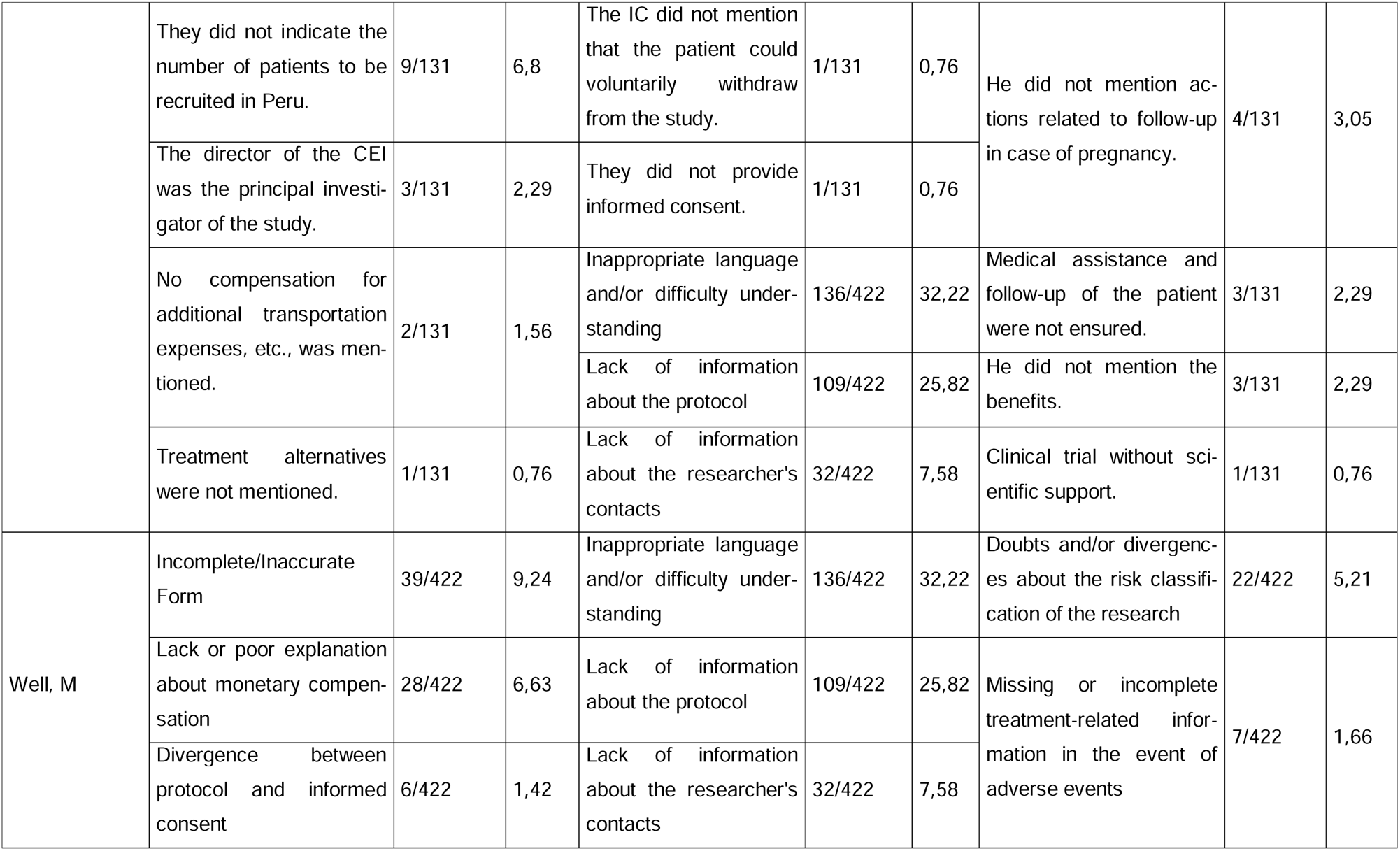
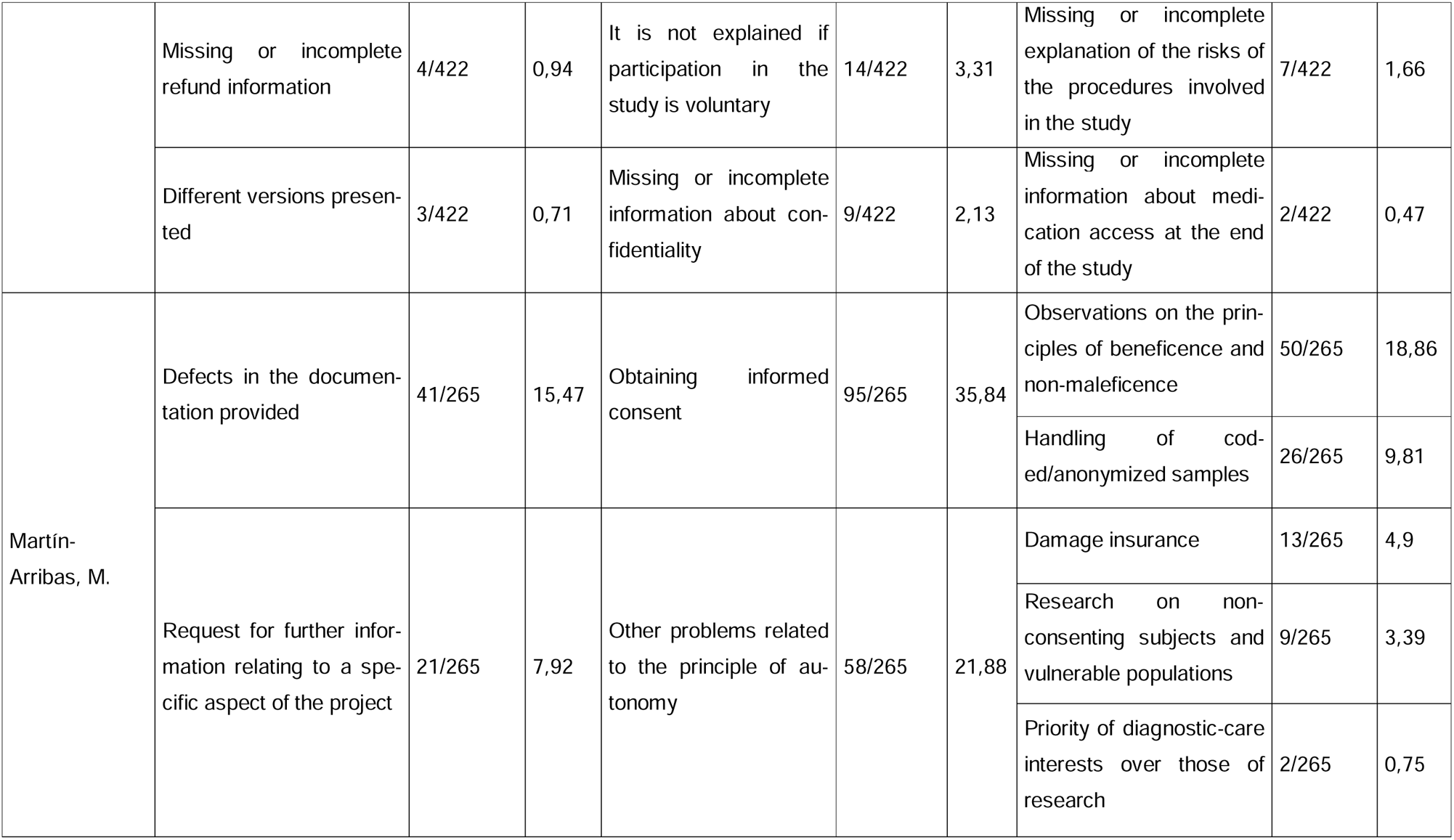
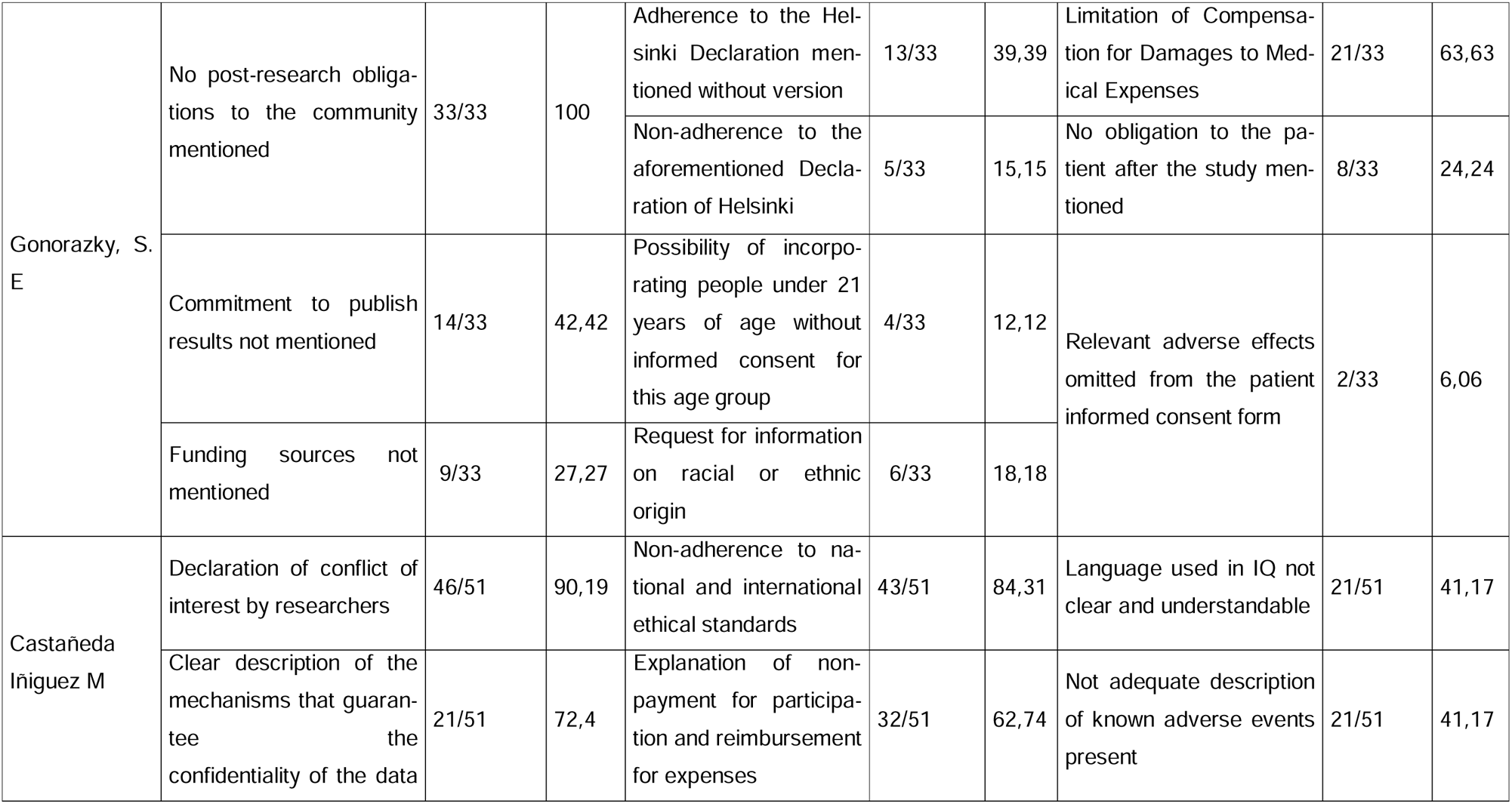
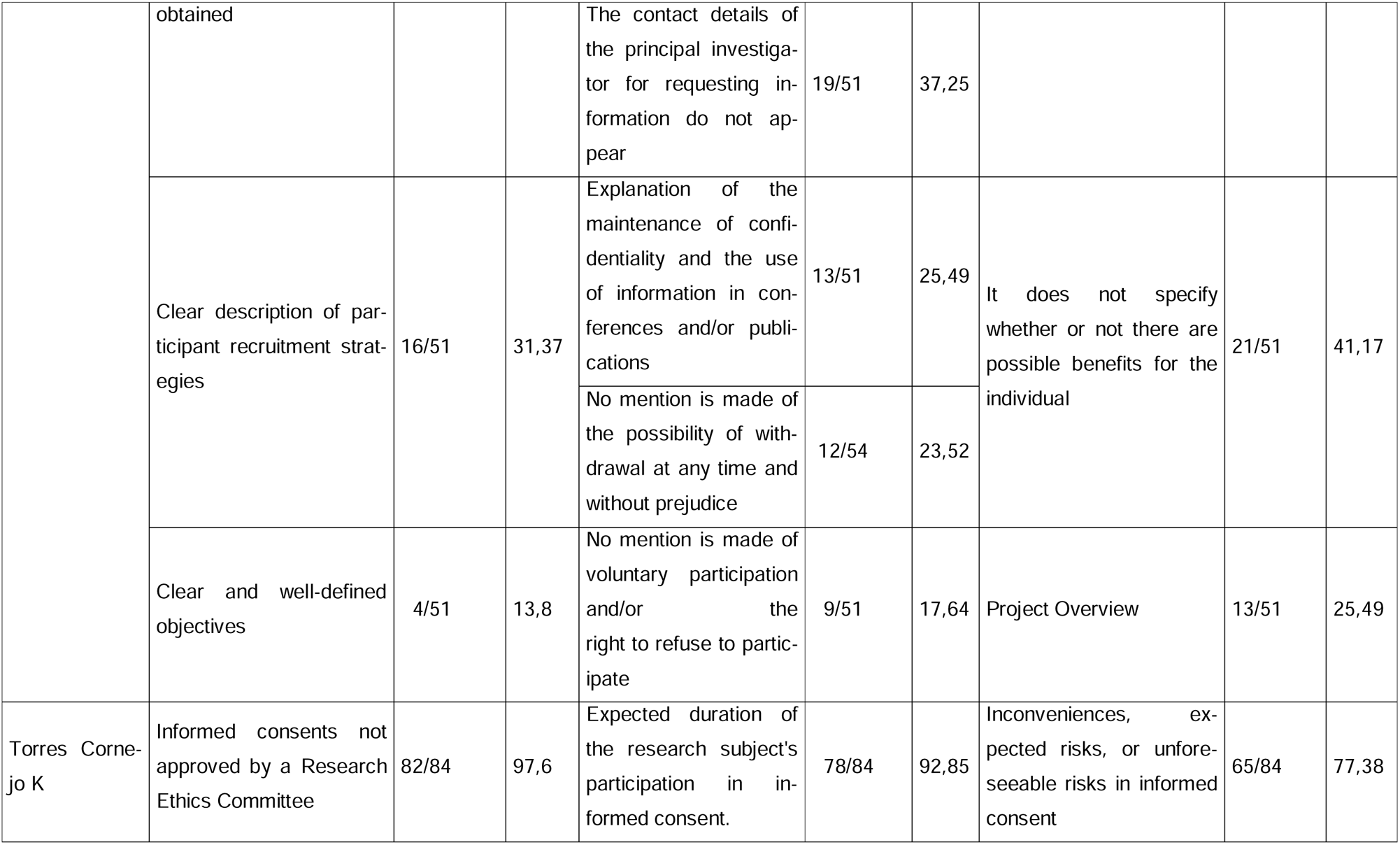
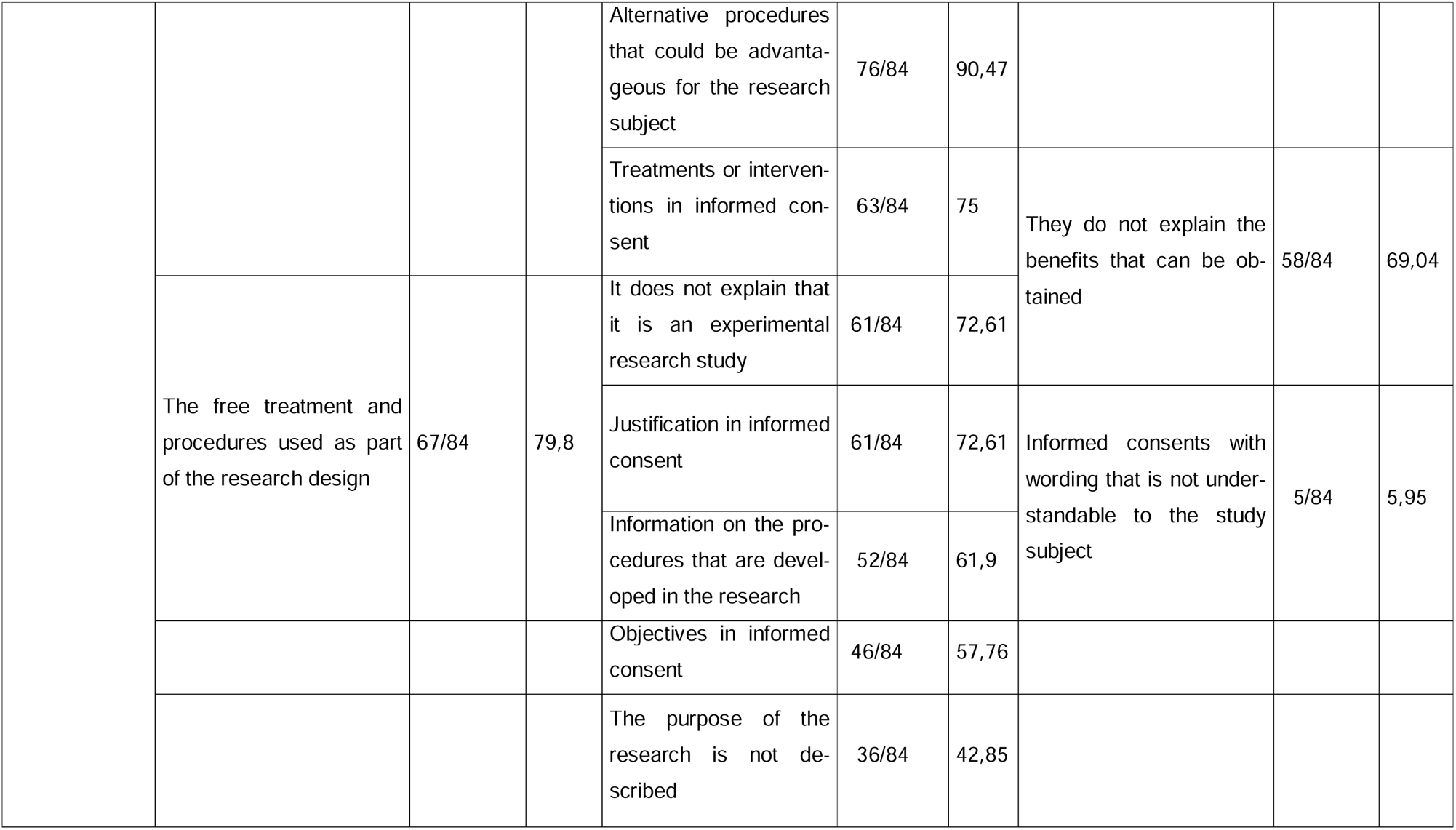
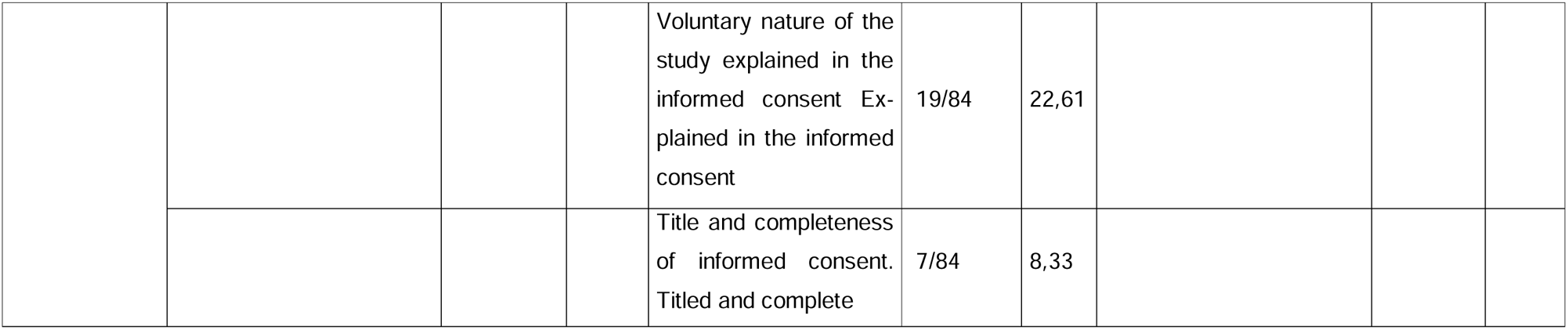
Observations on informed consent and ethical principles.

### 2. Results of the Meta-analysis

Considering the different ways in which data were presented in the included studies, two distinct meta-analyses were conducted. The first grouped the studies that reported the total number of observations per protocol evaluated, while the second meta-analysis was performed with data based on the number of protocols evaluated.

Given the detection of considerable heterogeneity, a subgroup analysis was carried out to identify patterns and differences in the results. Only those meta-analysis results whose heterogeneity was less than 75% are reported.

In the meta-analyses presented, the p-value obtained by Egger’s test was greater than 0.05.

#### a. Ethical Principle Justice

Figure 1 presents an analysis of studies that examined adherence to ethical principles in the protocols evaluated by the RECs. The results reveal that 97% (95% CI: 94-100) of the protocols received some request for amendment related to the ethical principle of justice.

**Figure 1.**
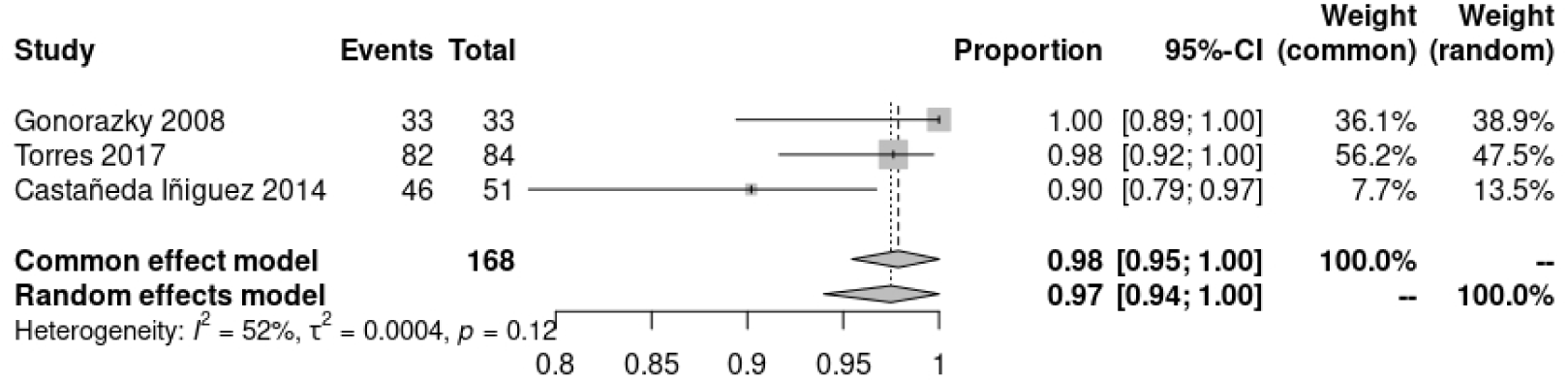
Meta-analysis according to the number of protocols evaluated.

Figure 2 presents the results of the meta-analysis, according to the number of observations per assessed protocol, revealing that in Latin America 9% (95% CI: 7-12) of observations related to the ethical principle of justice were found. In contrast, in Europe, this proportion rises to 15% (95% CI: 9-24), although there is notable heterogeneity among the studies analysed.

##### Metaanalysis according to the number of observations

**Figure 2.**
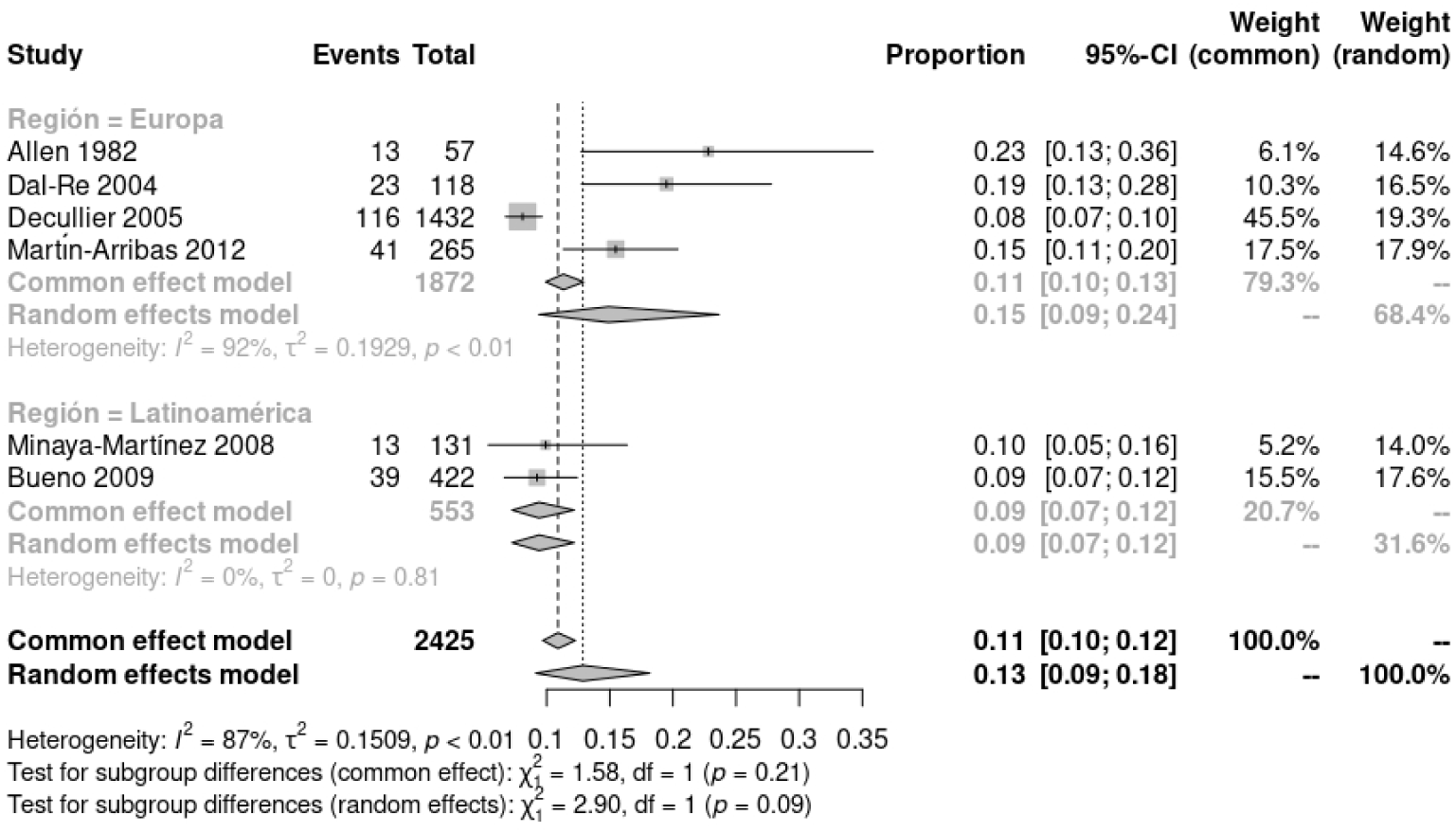
Metaanalysis of subgroups according to region.

#### b. Ethical principle autonomy/respect

When examining the number of requests for amendments by the number of protocols evaluated, it is observed that the proportion varies from 39.39% to 92.85%, (Heterogeneity = 88%) as reported by Castañeda Iñiguez et al. (2014), Gonorazky (2008) and Torres Cornejo (2017). (14–16)

The meta-analysis of Figure 3 indicates that the proportion of observations per protocol evaluated corresponds to 26% (95% CI 20, 33). When the protocol is specified as an experimental study, subgroup analysis indicates a proportion of 17% (95% CI 13, 22), while when the type of study is not specified or corresponds to observational studies, requests for amendments reach 31% (95% CI 27, 35).

**Figure 3.**
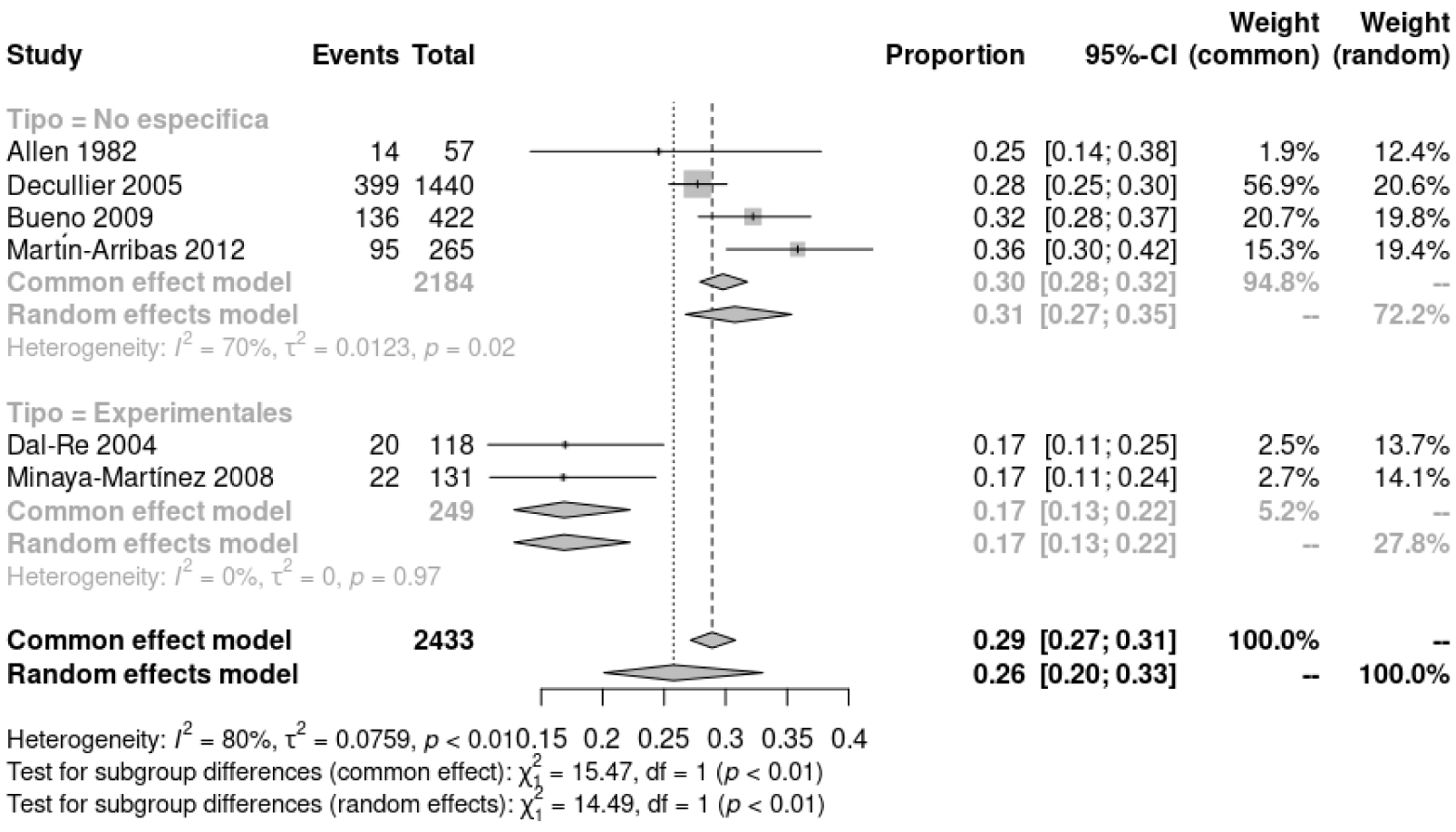
Metaanalysis of subgroups according to the type of protocol analyzed.

#### c. Ethical principle Beneficence

If we analyze the lack of adherence to the principle of beneficence according to the number of protocols evaluated, we have that the proportion ranges from 41.17% to 77.38% (heterogeneity = 85%) (17–19).

In relation to this ethical principle, objections regarding the number of observations made per protocol evaluated range from 5.26 % to 27.11 % (heterogeneity = 94%)(20,21).

Considering the exploratory nature of this study and that it does not seek to generate recommendations for clinical practice, the evaluation of the quality of the evidence according to the GRADE system is not included.

### 3. Narrative synthesis

Given that most of the meta-analyses showed a high heterogeneity, a narrative synthesis of the studies included in the present work is presented.

The Allen & Waters study examined the need to modify drug testing protocols and epidemiological studies in England from 1970 to 1981. It was found that approximately 33% of drug testing protocols and up to two-thirds of epidemiological studies required modifications between 1975 and 1979. During this period, the percentage of projects in need of adjustment increased markedly, from 10 per cent in 1971 to 39 per cent in 1979. (20)

Dal-Ré et al. examined 187 applications for multicenter trials evaluated by the Clinical Evaluation Ethics Committees in Spain between 2002 and 2003. Of the 183 applications approved, 101 were the subject of requests for clarification from 41 of the 62 participating IECs. The areas of improvement totaled 307 requests, with patient information being the most frequent aspect with 118 requests. Required changes included risk assessment, adverse events (27.1%), typographical and logistical aspects (19.5%), data protection (16.9%), procedures (11.9%), additional information (11.9%), clarity in terms (8.5%), and data deletion (4.2%) (21).

Decullier et al. evaluated 796 of the 1143 protocols approved by 25 of the 48 French IECs in 1994. 31% of the protocols were approved without modifications in an average of 16 days. Protocols with minor changes took 27 days and those requiring major changes took 48 days. 57% of the protocols needed clarification, and 19% required a second review. In total, 1438 points were cited for modification, with patient information (28%) and consent (18%) as the main reasons. 68% of the protocols investigated drugs, most of them were national (80%) and single-center studies. The workload was 24 hours per protocol, and 81% of the investigators did not send further information to the ECs (22)

Minaya-Martínez and Díaz-Sandoval analyzed the evaluation records of 91 clinical trials carried out by the Clinical Trials Committee (CEC) and previously approved by the corresponding RECs in Lima in 2006. We examined the ECC observations for each clinical trial, finding that only 11 received no observations. Of the 80 trials that did receive observations, 66% (53) had observations exclusively of an ethical nature. In total, 237 observations were registered, of which 131 were ethical, representing 55.3% of the total. However, the study does not provide information on the duration of the evaluations.(23)

The study by Bueno et al. evaluated 1,256 research projects submitted to the CEI of the Hospital das Clínicas of the University of São Paulo School of Medicine in 2007. The average evaluation time was 49.95 days. Most projects, 68% (857), were reviewed in a single meeting with an average of 39 days. Of the remainder, 27.2% (342) were reviewed in two meetings (average of 68.78 days), 4.2% (53) in three meetings (average of 99.62 days) and 0.3% (4) in four meetings (average of 127.30 days).

Of the 399 projects reviewed in multiple sessions, 357 were analyzed because they were research on human beings. Among these, 120 addressed special topics, and 1.4% (5) were rejected, while 1.1% (4) were submitted only for the committee’s knowledge. The main reasons for returning projects to researchers were problems with the informed consent form (52.7%) and inadequacies in the research protocol (25.6%), together representing 78.3% of the reasons for return. Other reasons included incomplete or incorrect documentation (8.5%), incomplete or incorrect registration form (8.1%), lack of timeline (2.6%), doubts about financial support (1.8%), and issues related to the *Comissão Nacional de Ética em Pesquisa* 0.5%. (24)

The study by Martín-Arribas et al. collected data from observations made on 100 protocols submitted to the Animal Ethics and Welfare Committee of the Carlos III Health Institute in Spain, between June 1, 2009 and June 30, 2010. A total of 265 observations were documented, classified into three categories: 76.5% (203) related to bioethics, 15.6% (41) due to poor documentation, and 7.9% (21) due to the need for additional project-specific information. The median of observations per project was 4 (mode 0; range 0-17), and the median of versions presented was 2 (1; 1-4). The evaluation process, from presentation to issuance of the favorable report, lasted an average of 13.5 days (median 13.5; mode 13; interval 1-95 days) (25)

The study by Gonorazky S analyzed 33 industry protocols presented at the Private Community Hospital in Argentina between 2005 and 2006, previously evaluated by a non-institutional CEI of national scope. 92 relevant objections were identified in 85% of the protocols. It found that 64% restricted compensation for damages to medical expenses arising from treatment, 15% made no mention of adherence to the Declaration of Helsinki, and 42% did not include a commitment to publish results. In addition, 24% allowed the study to be suspended without explanation and another 24% did not establish obligations towards patients at the end of the study. 27% did not mention the source of funding, 12% contemplated the inclusion of minors without informed consent, and 18% requested sensitive data such as racial or ethnic origin. 6% omitted relevant adverse effects in informed consent, and none mentioned post-research obligations to the community. The study does not specify the type of studies analyzed (14).

The study by Castañeda Iñiguez et al. evaluated the application of ethical guidelines in medical research projects funded by the National Commission for Scientific and Technological Research in Chile between 2006 and 2008. 51 projects were analyzed, 22 financed by the National Fund for Research and Development in Health (FONIS) and 29 by the National Fund for Scientific and Technological Development (FONDECYT). It was found that 68.2% of the FONIS projects and 72.4% of the FONDECYT projects lacked a clear description of the mechanisms to guarantee the confidentiality of the data. In addition, 63.6% and 69% of the projects, respectively, did not address the potential risks to the participants, and only 31.8% and 34.5% mentioned the benefits to the participants. Only 18.2% of the FONIS projects and 13.8% of the FONDECYT projects complied with national and international ethical standards in the drafting of informed consent. Conflict of interest reporting was minimal in both groups, and the study noted that 60% of the principal investigators were male (15).

Torres Cornejo’s study evaluated the quality of the informed consent form in 84 graduate theses from the Catholic University of Santa María de Arequipa, Peru, between January 2013 and December 2017. It was found that 97.6% of the informed consent forms were not approved by an IEC. Only 71.4% included the full degree and 77.4% explained that the study was voluntary. 72.6% did not clearly explain what the study was about, 60.7% did not present justification and 54.8% did not mention the objective of the study. In addition, 75% did not describe the treatments or interventions, 61.9% did not include the procedure to be used, and 92.9% did not indicate the expected duration of participation. 79.8% did not mention the free treatment and procedures, and 69% did not describe the expected benefits. Finally, 90.5% did not describe alternative procedures that could be advantageous for the research subjects. (16)

## Discussion

This study provides data on the work of the RECs in the evaluation of research protocols, as well as the adherence of research protocols to established ethical principles.

In the meta-analyses, a significant disparity in the number of observations between different regions was identified, raising questions about the possible causes or factors influencing this variation in the ethical evaluation of research protocols. This discrepancy could be attributed to differences in the interpretation of ethical principles, cultural approaches to research, or even disparities in institutional resources and capacities (20–24).

There is also a difference in the number of observations between observational and non-observational (experimental) studies. This disparity can be explained by the inherent characteristics of each type of research and the associated ethical risks. Experimental studies, by involving manipulation of variables and random assignment of treatments, may carry additional risks for participants, making them more likely to be scrutinized closely by Ethics Committees, as well as to be led by more experienced researchers. On the other hand, observational studies, while they may be less intrusive, are not without ethical concerns, such as data privacy and confidentiality. The protocols of these studies should also be carefully developed and presented, but they might require fewer ethical observations compared to experimental ones(20–25).

### a. Justice

In the present work, it was found that all the study protocols had some objection related to the principle of justice. In addition, it was noted that more observations are made in Europe in relation to this principle compared to other regions. It is relevant to note that the IECs in Europe have not been considered negligent as has happened in the United States and Canada. Justice focuses on the societal benefit derived from knowledge gained through research and on the fair selection of participants for equitable outcomes. This principle does not directly relate to the application of sample-size formulas, but rather involves broader considerations about equity in research. The principle of justice is fundamental in the ethics of medical research, ensuring an equitable distribution of benefits and risks at both the individual and societal levels. This principle guides the impartial selection of research subjects, avoiding unfair privileges and protecting vulnerable people from additional burdens. In addition, research processes and documents must be culturally and linguistically adapted to increase equity. Researchers must rigorously evaluate the risk-benefit ratio and protect subjects from conditions that may increase their vulnerability. This equity strengthens public trust and ensures that scientific advances fairly benefit society as a whole, given that much research is carried out with public funds (4,26–29)

### b. Principle of autonomy

The total proportion of observations found in the present work corresponds to 26% and when it is specified that the protocol corresponds to an experimental study it corresponds to 17%, while when the type of study is not specified or it is observational it reaches 31%. When examining the number of protocols evaluated, the proportion ranges from 39.39% to 92.85%. Experimental studies, despite their greater complexity, received fewer objections. However, this can be attributed to several factors. First, these studies are usually led by experienced researchers with adequate resources, who comply with high ethical and regulatory standards from the outset, anticipating objections and preparing detailed protocols. Second, due to their nature, experimental studies face rigorous pre-approval reviews, which corrects ethical issues before reaching RECs. Third, the institutions that conduct these studies implement internal committees and ongoing oversight to ensure ethical compliance, decreasing the need for additional observations by the IRBs (4,5,30,31).

The principle of autonomy focuses on respect for people’s ability to freely decide on their participation in studies. This materialized through informed consent, where participants must receive detailed information about the objectives, procedures and risks of the study, and give their consent or refuse to participate freely and voluntarily, with the possibility of withdrawing at any time. It is especially crucial in the protection of vulnerable populations, such as children or patients with reduced mental capacities, who require additional measures to ensure their autonomous participation or authorized by a legal representative. In addition, respect for autonomy implies safeguarding the privacy and confidentiality of the personal information provided by the participants. The confidentiality of personal information and the responsible use of biological samples must be guaranteed. All these principles should be reviewed by research ethics committees to ensure adequate compliance with these principles, including studies with minimal risk or risk-free research (5,9,32–36).

### b. Beneficence

Objections regarding the number of observations made to the protocols range from 5.26% to 27.11%. If we analyze the lack of adherence to the principle of beneficence according to the number of protocols evaluated, we have that the proportion ranges from 41.17% to 77.38%. The principles of beneficence and non-maleficence are closely linked and state that research should prioritize the well-being of participants, maximizing potential benefits and minimizing risks. In studies involving vulnerable populations or people without the capacity to consent, these ethical considerations are especially relevant. It is crucial that the well-being of the participants is protected above the objectives of the research, implementing measures to avoid any harm. Traditionally, it was believed that the role of IECs was only to safeguard the dignity, rights, safety, and well-being of participants. However, it is now recognised that RECs must also assess whether the proposed research promotes scientific validity, analysing the balance between the study’s risk and its potential benefit to both the current population and future generations (4,5,7,9).

Ultimately, RECs play a crucial role in biomedical research, balancing the value of scientific knowledge with the protection of participants. Its main function is to safeguard the safety and dignity of research subjects, offering an independent, impartial and timely evaluation. RECs should be composed of trained members, to properly understand research proposals and provide clear and timely responses to researchers. (4)

In subgroup analyses, some results are likely to be significant by chance, although they may also be influenced by cultural differences in research between the regions studied. This study revealed high heterogeneity in the meta-analyses, attributable to the various local laws, although the ethical principles are universal. The Egger test applied only in groups with less than 75% heterogeneity suggests absence of publication bias, although it should be interpreted with caution due to the limited number and small sample sizes included, mostly case series. Despite these limitations, this study offers the best evidence available to date in an observational exploratory context, consistent with previous research on the evolution and ethical variation in medical research(4,5,36).

## Conclusion

The proportions of non-adherence to ethical principles by the CEIs were determined, where it was found that the ethical principle of justice is the one with the most observations according to the number of protocols evaluated. This study highlights the variability in the observations made by the RECs on medical research protocols in terms of adherence to the ethical principles of autonomy, beneficence, and justice. The differences observed between regions and types of studies underscore the influence of cultural, interpretive, and institutional factors on ethical evaluation. Experimental studies, despite their greater complexity, received fewer observations. The findings indicate the need to strengthen the resources and capacities of RECs to ensure thorough and equitable ethical assessments. It is essential to promote a fair selection of research subjects, respect their autonomy, and maximize benefits to the population while minimizing risks to study subjects.

## Data Availability

All data produced in the present work are contained in the manuscript

## Recommendations

It is recommended that IRCs publish the results of their activities to increase the transparency of their work. This will not only provide useful information for researchers but will also help motivate them in their research work.

## Financing

The present work, as well as the previous research, have not received any funding for its elaboration.

